# Radiomics-based prediction of local control in patients with brain metastases following postoperative stereotactic radiotherapy

**DOI:** 10.1101/2024.01.03.24300782

**Authors:** Josef A. Buchner, Florian Kofler, Michael Mayinger, Sebastian M. Christ, Thomas B. Brunner, Andrea Wittig, Bjoern Menze, Claus Zimmer, Bernhard Meyer, Matthias Guckenberger, Nicolaus Andratschke, Rami A. El Shafie, Jürgen Debus, Susanne Rogers, Oliver Riesterer, Katrin Schulze, Horst J. Feldmann, Oliver Blanck, Constantinos Zamboglou, Konstantinos Ferentinos, Angelika Bilger-Zähringer, Anca L. Grosu, Robert Wolff, Marie Piraud, Kerstin A. Eitz, Stephanie E. Combs, Denise Bernhardt, Daniel Rueckert, Benedikt Wiestler, Jan C. Peeken

## Abstract

**Background:** Surgical resection is the standard of care for patients with large or symptomatic brain metastases (BMs). Despite improved local control after adjuvant stereotactic radiotherapy, the local failure (LF) risk persists. Therefore, we aimed to develop and externally validate a pre-therapeutic radiomics-based prediction tool to identify patients at high LF risk.

**Methods:** Data were collected from *A Multicenter Analysis of Stereotactic Radiotherapy to the Resection Cavity of Brain Metastases* (AURORA) retrospective study (training cohort: 253 patients (two centers); external test cohort: 99 patients (five centers)). Radiomic features were extracted from the contrast-enhancing BM (T1-CE MRI sequence) and the surrounding edema (FLAIR sequence). Different combinations of radiomic and clinical features were compared. The final models were trained on the entire training cohort with the best parameters previously determined by internal 5-fold cross-validation and tested on the external test set.

**Results:** The best performance in the external test was achieved by an elastic net regression model trained with a combination of radiomic and clinical features with a concordance index (CI) of 0.77, outperforming any clinical model (best CI: 0.70). The model effectively stratified patients by LF risk in a Kaplan-Meier analysis (p < 0.001) and demonstrated an incremental net clinical benefit. At 24 months, we found LF in 9% and 74% of the low and high-risk groups, respectively.

**Conclusions:** A combination of clinical and radiomic features predicted freedom from LF better than any clinical feature set alone. Patients at high risk for LF may benefit from stricter follow-up routines or intensified therapy.

**Key points:**

- Radiomics can predict the freedom from local failure in brain metastasis patients
- Clinical and MRI-based radiomic features combined performed better than either alone
- The proposed model significantly stratifies patients according to their risk

**Importance of the Study:** Local failure after treatment of brain metastases has a severe impact on patients, often resulting in additional therapy and loss of quality of life. This multicenter study investigated the possibility of predicting local failure of brain metastases after surgical resection and stereotactic radiotherapy using radiomic features extracted from the contrast-enhancing metastases and the surrounding FLAIR-hyperintense edema.

By interpreting this as a survival task rather than a classification task, we were able to predict the freedom from failure probability at different time points and appropriately account for the censoring present in clinical time-to-event data.

We found that synergistically combining clinical and imaging data performed better than either alone in the multicenter external test cohort, highlighting the potential of multimodal data analysis in this challenging task. Our results could improve the management of patients with brain metastases by tailoring follow-up and therapy to their individual risk of local failure.

## Introduction

Brain metastases (BMs) are the most common malignant brain tumors, far outnumbering primary brain tumors such as gliomas^1^. Recent guidelines recommend surgery as a treatment for patients with symptomatic or large BMs^2^. To improve local control, stereotactic radiotherapy (SRT) should be applied to the resection cavity in patients with one to two resected BMs^2^. This way, local control rates of 70% to 90% at twelve months can be achieved^3^.

Determining an individual patient’s risk of local recurrence can benefit patients by tailoring follow-up treatment and care. For example, patients at high risk of local failure may benefit from SRT dose escalation, systemic therapy agents with penetration of the blood-brain barrier, and more frequent follow-up imaging after SRT to detect a potential failure early.

Recent publications have demonstrated the power of automated segmentation of BMs and their surrounding edema^4–6^. This cannot only help radiation oncologists by eliminating the time-consuming task of manual BM delineation but can also simplify other additional evaluations: Radiomics allows the extraction of large amounts of quantitative imaging features from a previously delineated image^7^. This enables professionals to analyze additional information that is not visible to the human eye and allows the creation of predictive mathematical models^8^.

Such radiomics models can be used for multiple tasks such as tumor characterization, prediction of treatment response, and prognostic risk assessment^9–13^.

Some radiomic features are sensitive to acquisition modes and reconstruction parameters^14^. Furthermore, MRI intensities are not standardized and depend on the manufacturer and model of the devices^15^. Moreover, patients and treatment characteristics can differ between medical institutions. Therefore, multicentric training and testing are needed to develop and validate generalizable models.

Several previous studies could demonstrate the general propensity of radiomics to predict local failure (LF) as a binary variable in patients receiving stereotactic radiotherapy without surgery in monocentric studies without external validation^16–18^.

The aim of this project was to develop a pre-therapeutic radiomics-based machine learning model to predict freedom from local failure (FFLF) after surgical resection and SRT of BMs. All models were validated in an external multicenter international test cohort. The ability to stratify patients into specific risk groups and their net clinical benefit were assessed.

## Methods

### AURORA study

MR imaging and clinical data was collected as part of the “*A Multicenter Analysis of Stereotactic Radiotherapy to the Resection Cavity of Brain Metastases”* (AURORA) retrospective trial. The trial was supported by the *Radiosurgery and Stereotactic Radiotherapy Working Group* of the *German Society for Radiation Oncology* (DEGRO). The inclusion criteria were: known primary tumor with resected BM and SRT with a radiation dose of > 5 Gy per fraction. Exclusion criteria were: interval between surgery and RT > 100 days, premature discontinuation of RT, and any previous cranial radiation therapy (RT). Synchronous non-resected BMs had to be treated simultaneously with SRT. Ethical approval was obtained at each institution (main approval at the Technical University of Munich: 119/19 S-SR).

LF was determined by individual radiologic review or by histologic results after recurrence surgery. FFLF was calculated as the time difference between the end of SRT and LF. If no LF occurred, patients were right-censored after the last available imaging follow-up.

### Dataset

In total, we collected data from 481 patients from seven centers. We analyzed four preoperative imaging sequences of each patient: a T1-weighted sequence with and without contrast enhancement (T1-CE and T1), a T2-weighted sequence (T2) as well as a T2 fluid-attenuated inversion recovery sequence (T2-FLAIR). Except for T1-CE, a missing sequence was allowed.

The required data were available for 352 patients. We split the patients into a training cohort with 253 patients from two centers and an external, multicenter, international test cohort with 99 patients from five centers.

### Preprocessing

The DICOM (Digital Imaging and Communications in Medicine file format) images were converted to NIfTI (Neuroimaging Informatics Technology Initiative file format) using dcm2niix^19^. The MRI sequences were then further preprocessed using BraTS-Toolkit^20^. First, the sequences were co-registered using niftyreg^21^ and these were then transformed into the T1-CE space. A brain mask was created using HD-BET^22^ and applied to all sequences to extract only the brain without the surrounding skull. The skull-stripped sequences were transformed into the BraTS space using the SRI-24 atlas^23^. Overall, the preprocessing provided co-registered, skull-stripped sequences in a 1 millimeter isotropic resolution in BraTS space.

The missing sequences were then synthesized using a generative adversarial network (GAN). The GAN takes the three available sequences as input and generates the matching missing fourth sequence. We used a GAN which was originally developed for missing sequences in glioma imaging^24^, but has been proven to work for metastasis imaging^4,5^.

### Segmentation

All contrast-enhancing metastases and their surrounding edema were individually segmented using the open-source software 3D-Slicer (version 4.13.0, stable release, https://www.slicer.org/)^25^ by a medical doctoral student (JAB) after undergoing extensive training by a board-certified radiation oncologist (JCP) (7 years of experience). To ensure accuracy, all segmentations for the test cohort were reviewed and manually adjusted by JCP.

To test the feasibility of a fully automated workflow, segmentations generated by a previously trained neural network^4,5^ were used as alternative segmentations and compared to the manual segmentations.

As around 25% of patients had multiple BMs, but usually only the largest is resected^26^, we also determined the largest metastasis with a connected component analysis^27^ in all patients with multiple BMs and used only that metastasis and its surrounding edema as segmentations for an additional analysis.

### Radiomic feature extraction

Radiomic features were extracted with pyradiomics (version 3.0.1, https://github.com/AIM-Harvard/pyradiomics)^28^ using the Python implementation. The metastasis segmentation was used to extract the T1-CE features, while the edema segmentation was used for the T2-FLAIR features. In total, we extracted 104 original features per segmentation (see Supplemental Table 1 for a list of features and extraction parameters).

Further analysis and modeling were performed in the programming language R 4.2.3^29^. To adhere to the *Image Biomarker Standardisation Initiative* (IBSI) standard^30^, the kurtosis was adjusted by −3. We created nine feature sets in total. Three of these included only radiomic features. The *metastasis* and *edema* feature sets were created by extracting the features from the T1-CE sequence and T2-FLAIR sequence, respectively. Both feature sets were merged into a *combined* feature set. We also created three clinical feature sets with the following clinical features:

- *pre-OP* feature set: patient age at RT start, Karnofsky performance status (KPS), histology of the primary tumor, location of BM
- *post-OP* feature set: *pre-OP* + resection status
- *RT* feature set: *post-OP* + concurrent chemotherapy, concurrent immunotherapy and equivalent dose in two Gray fractions (EQD2)

As a seventh feature set, we combined all radiomic features (*combined*) with the *pre-OP* feature set to *comb+pre-OP*.

Multiple publications suggest the predictive value of the brain metastasis volume (BMV) for predicting LF^31–33^. Therefore, we created two additional feature sets by adding the cumulative BMV of each patient as an additional feature to the *pre-OP* set (*pre-OP+BMV*) and the *comb+pre-OP* set (*comb+pre-OP+BMV*).

### Intraclass correlation

To identify radiomic features that were susceptible to small changes in segmentation, we generated additional segmentations of all patients in the training cohort using the previously mentioned neural network^4^. Intraclass correlation (ICC (3,1)) was calculated using the R package “irr”^34^. According to Koo *et al*., an ICC above 0.75 is considered “good”^35^. Consequently, this value was employed as a cut-off threshold. Of the 208 features, 173 (83%) had an ICC of > 0.75 and were selected for all further steps. Of the 35 excluded features, the majority (27) were extracted from the edema mask, while only eight excluded features were extracted from the metastasis mask.

All selected radiomic features were normalized by z-score standardization and by applying the Yeo-Johnson transformation^36^ to transform the distribution of a variable into a Gaussian distribution.

### Feature reduction

We applied a minimum redundancy - maximum relevance (MRMR) ensemble feature selection framework implemented in R^37^ initially proposed by Ding *et al*.^38^ as an efficient method for the selection of relevant and non-redundant features.

We created multiple smaller feature sets of the *metastasis*, *edema*, and *combined* feature sets with three, five, seven, nine, eleven, thirteen, and fifteen features each.

We used bootstrapping^39^ to obtain more reliable results: Feature reduction was repeatedly applied to 1000 bootstrap samples for each set and each number of features. For our final set of features, we ranked the features based on the number of times they were selected.

The best number of features was later determined by nested cross-validation in the training set.

### Batch harmonization

To account for differences created by 29 different MRI scanners in our multicenter dataset, we used batch harmonization implemented by neuroCombat^40^. In total, 10 batches were created according to the MRI model names by combining related models. According to Leithner *et al*.^41^, ComBat harmonization without Empirical Bayes estimation provided slightly higher performance in similar machine learning tasks. Therefore, Empirical Bayes was deactivated. Besides the non-harmonized dataset, we created two harmonized datasets: one by only adjusting the means and the other by adjusting means and variances.

### Model creation, testing, and patient stratification

For model creation and evaluation, the R package MLR3^42^ was used as a basis. Our prediction target was right-censored time-to-event data, where we used LF as the event and the FFLF or time-to-last imaging follow-up as the time variable for patients with and without event, respectively. We compared three different learners: random forest (RF), extreme gradient boosting (xgboost), and generalized linear models with elastic net regularization learner (ENR).

We implemented nested cross-validation to select the best mode of batch harmonization and the best number of features: For batch harmonization selection, all three datasets were compared while always using the *combined* feature set with nine features. Five iterations of five-fold nested cross-validation for dataset selection showed no significant difference between the sets with and without batch harmonization (p = 0.3, Kruskal-Wallis rank sum test). Therefore, all further analyses were performed on the base dataset without batch harmonization to avoid unnecessary and potentially distorting preprocessing steps. To select the ideal number of features in each feature set, the nested cross-validation was conducted without batch harmonization. The best average performance was achieved with seven, three, and seven features in the *metastasis*, *edema*, and *combined* sets, respectively. The *comb+pre-OP* set, which included the seven *combined* and four *clinical* features, therefore, had 11 features. The features are listed in Supplementary Table 2.

The parameter tuning was performed using repeated cross-validation. All tuning and selection steps were performed on the training set. To account for the class imbalance (around 1:5 event:no-event), synthetic minority over-sampling was implemented using SMOTE^43^. We used an implementation in R which is capable of handling numeric and categorical data. The number of samples in the minority class was increased by creating synthetic samples to reach a ratio of 1:2. We only used SMOTE on the training folds in each step of our (nested) cross-validation. This way we ensured that our models were only validated on real patients.

The final models were trained with the best parameters determined by the cross-validation on the whole training set while also using SMOTE to balance the classes. The models were then tested on our multicentric external test cohort.

The 33rd and 66th percentiles of the continuous risk ranks in the training cohort were used as cutoffs for patient stratification. These cutoffs were used to divide the test cohort into three groups according to their predicted continuous risk rank and compare their survival with Kaplan Meier analysis.

### Metrics

To account for both timing and outcome, the learners’ performance was quantified using the concordance index (CI)^44^. The 95% confidence intervals are based on 10,000 bootstrap samples. A decision curve analysis was performed to consider clinical consequences with a time endpoint of 24 months^45^. The threshold range was chosen as suggested by Vickers *et al.*^46^ based on these considerations: Since LF is a severe event and its detection is critical, a lower threshold of 5% seems appropriate. Especially in elderly and multimorbid patients, where additional imaging may be burdensome, an upper threshold of 30% is reasonable.

## Results

An overview of patient characteristics of both patient cohorts is shown in Table 1. A total of 147 patients had missing sequences, the majority of which were missing T2 and T1 sequences (82% and 10%, respectively), which were not relevant for our further analyses. The general workflow, with example images of a test cohort patient, is shown in Figure 1.

**Figure 1:**
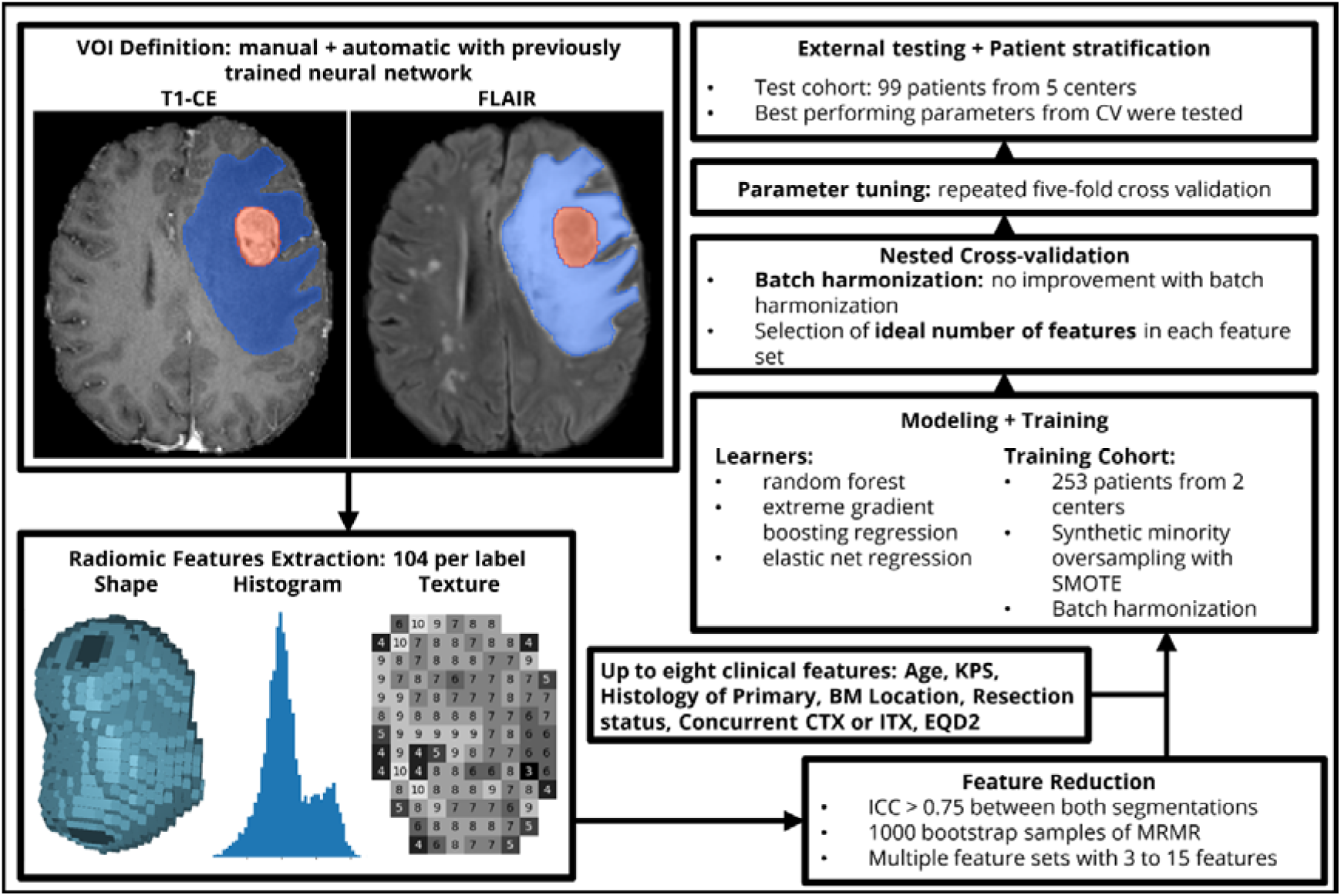
Summarized overview of our workflow. After manual and automatic definition of the volume of interest (VOI), we extracted 104 original features from each metastasis and edema segmentation. We reduced the number of features in each set with MRMR. Furthermore, we added up to eight clinical features and combined all features into multiple different feature sets. The optimal number of features in each set was determined with a nested cross-validation. The optimal parameters for our selected learners were chosen based on a 5-fold cross-validation. The best parameters for each learner-feature-combination were tested in the external test cohort.

**Table 1:**
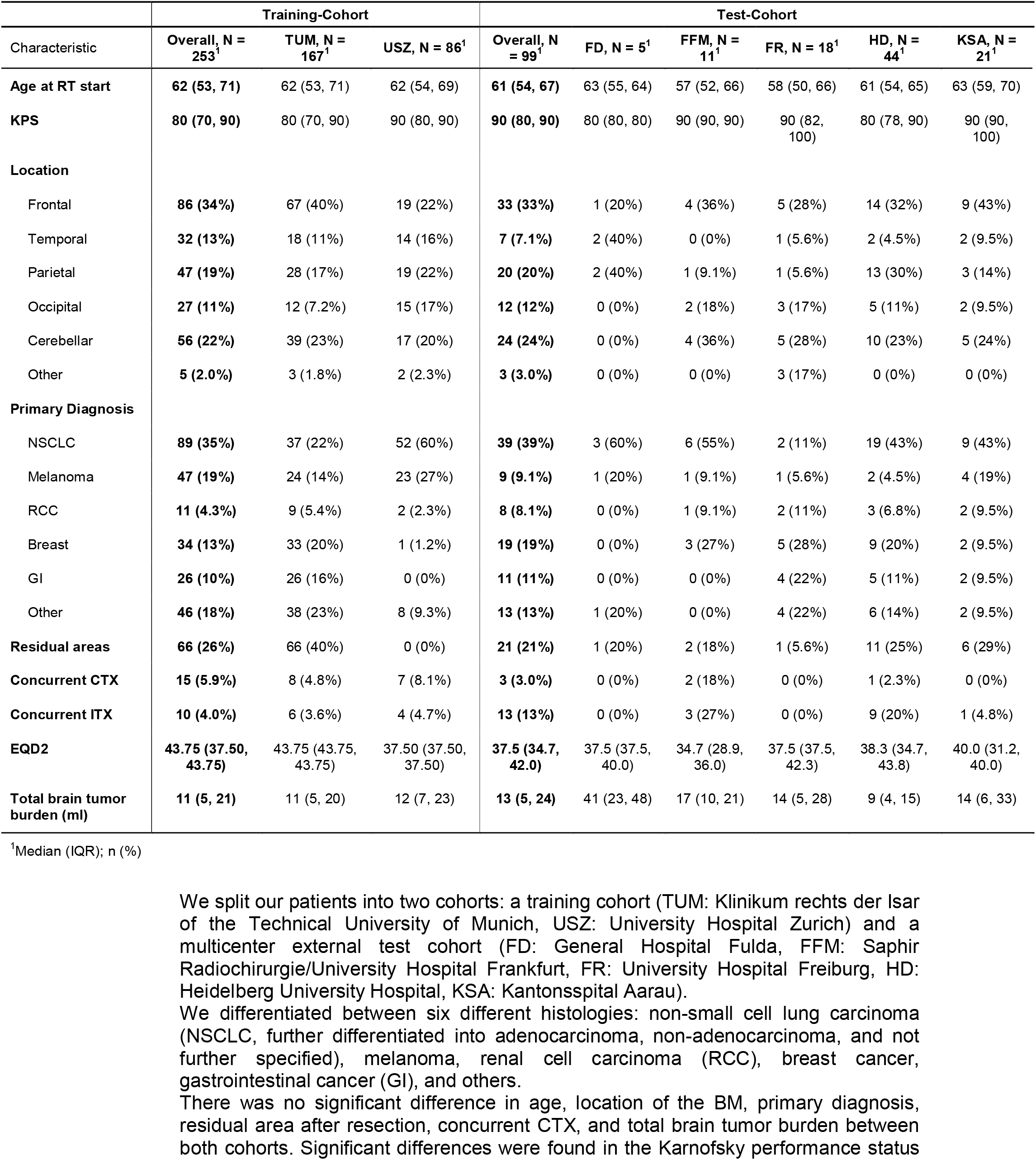
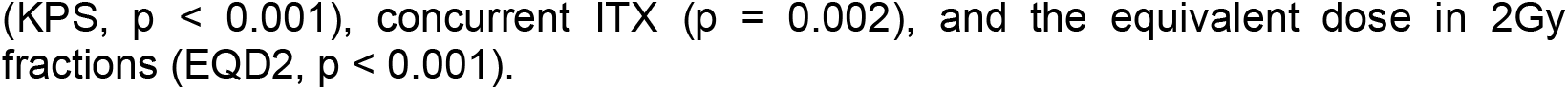
Cohort demographics.

### Baseline clinical models

To create a baseline for comparison with our radiomic models, we first tested the predictive value of two established clinical indices with univariate Cox analysis: the Recursive Partitioning Analysis (RPA)^47^ and the Graded Prognostic Assessment (GPA)^48^ index. They reached a CI of 0.47 and 0.52 in the internal validation, respectively. In external testing, RPA again performed worse with a CI of 0.39 compared to GPA with a CI of 0.44.

### Model performance

The performances in the internal validation, as well as in the multicentric external test cohort, are shown in Table 2. To determine the best overall learner, we ranked the performance across all feature sets and found that ENR ranked best, followed by RF and xgboost with mean ranks of 1.4, 1.6, and 2.9, respectively. Therefore, all further experiments were conducted with ENR. For completeness, the results obtained by RF and xgboost are shown in Supplementary Tables 3 and 4. The highest mean CI across all five folds and ten iterations of the cross-validation was achieved with the *comb+pre-OP* feature set (CI = 0.67).

**Table 2:** Performance in internal validation and external testing.

| Group | Learner | pre-OP | pre-OP + BMV | Post-OP | RT | T1-CE | FLAIR | comb | comb + pre-OP | comb + pre-OP + BMV |
| --- | --- | --- | --- | --- | --- | --- | --- | --- | --- | --- |
| 5-fold CV | ENR | 0.64 | 0.63 | 0.63 | 0.63 | 0.65 | 0.47 | 0.62 | **0.67** | 0.67 |
|  | RF | 0.63 | 0.63 | 0.63 | 0.63 | 0.61 | 0.58 | 0.64 | 0.66 | 0.66 |
|  | xgboost | 0.54 | 0.56 | 0.53 | 0.56 | 0.58 | 0.55 | 0.62 | 0.65 | 0.64 |
| external test cohort | ENR | 0.70 | 0.70 | 0.65 | 0.70 | 0.76 | 0.50 | 0.69 | **0.77** | 0.72 |
|  |  | (0.53-0.83) | (0.54-0.83) | (0.51-0.82) | (0.56-0.83) | (0.63-0.84) | (NA-NA) | (0.55-0.80) | (0.61-0.87) | (0.57-0.82) |

Parameter tuning and internal validation were performed with ten iterations of a five-fold cross-validation. The 95% confidence intervals (in parenthesis) are based on 10000 bootstrap samples. The combination of ENR learner and *comb+pre-OP* feature set performed best with a mean CI of 0.67. Adding BMV did not improve performance. By ranking the performance of the models across all feature sets, we identified ENR as the best learner and, therefore, tested this learner on the external test cohort. Again, the best performance was seen with the *comb+pre-OP* feature set (CI = 0.77). Best performance is printed in bold for the internal and external cohort.

The *comb+pre-OP* feature set also led to the highest performance in the external test cohort and achieved a CI of 0.77. While the *T1-CE* feature set achieved a CI of 0.76, *FLAIR* was only able to reach 0.50. The three clinical feature sets performed slightly worse than our radiomic feature sets or the combined feature sets: the *pre-OP*, *post-OP*, and *RT* feature sets reached a CI of 0.64, 0.63, and 0.63 in the internal validation, respectively. In external testing, they achieved a CI of 0.70, 0.65, and 0.70, respectively. While adding the BMV to the *pre-OP* feature set did not change the predictive performance, adding it to *comb+pre-OP* led to worse results with a CI of 0.72.

For reproducibility, we listed the beta values used by our best model (*comb+pre-OP* ENR) in Supplementary Table 5. The corresponding calibration curve to this model is shown in Figure 2 (right panel). Furthermore, we calculated the time-dependent area under the receiver operating characteristic curve (AUC) by transforming the crank to an event probability distribution. The proposed model reached a mean of 0.80. Supplementary Figure 1 shows the plotted time-dependent AUC.

**Figure 2:**
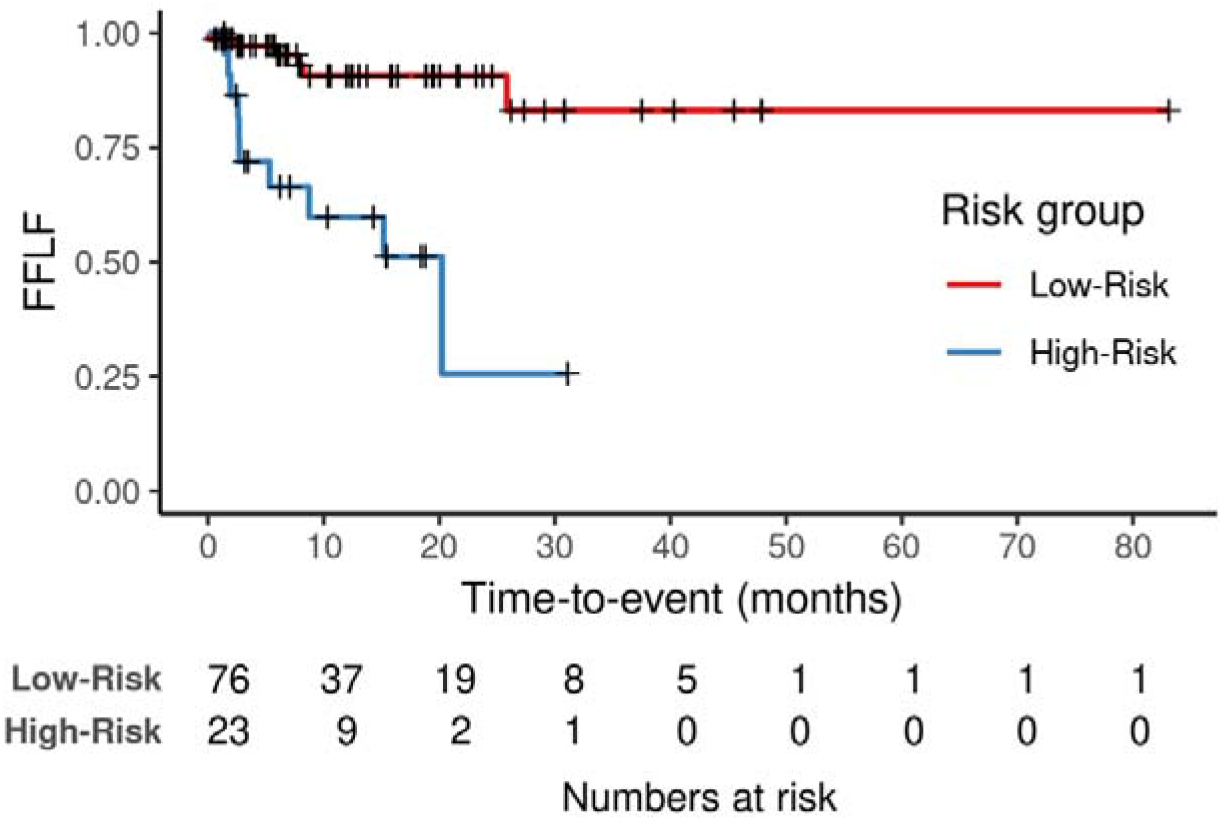
Kaplan Meier analysis. We created dichotomous predictions of the *comb+pre-OP* ENR model by using the 66th percentiles of the continuous risk ranks in the training cohort as cutoffs for patient stratification. We found a significant difference in freedom from local failure (FFLF) between the predicted low- and high-risk groups (p < 0.001) in the multicenter external test cohort. After 24 months, we found a FFLF of 91% and 26% in the groups, respectively.

### Patient stratification

Using the cutoffs determined by the training cohort as described above, our *comb+pre-OP* ENR model was able to significantly stratify the patients into three risk groups with a low, medium, and high risk of local failure (p = 0.0001, Chi-squared Test). A Kaplan-Meier analysis with all three groups is shown in Supplementary Figure 2.

By combining the low- and medium-risk groups into one, we created dichotomous predictions. Kaplan-Meier analysis (Figure 2) illustrates the survival in each risk group. Decision curve analysis using these predictions showed a net benefit of our predictive model compared to treating all patients in the relevant threshold range (Figure 3).

**Figure 3:**
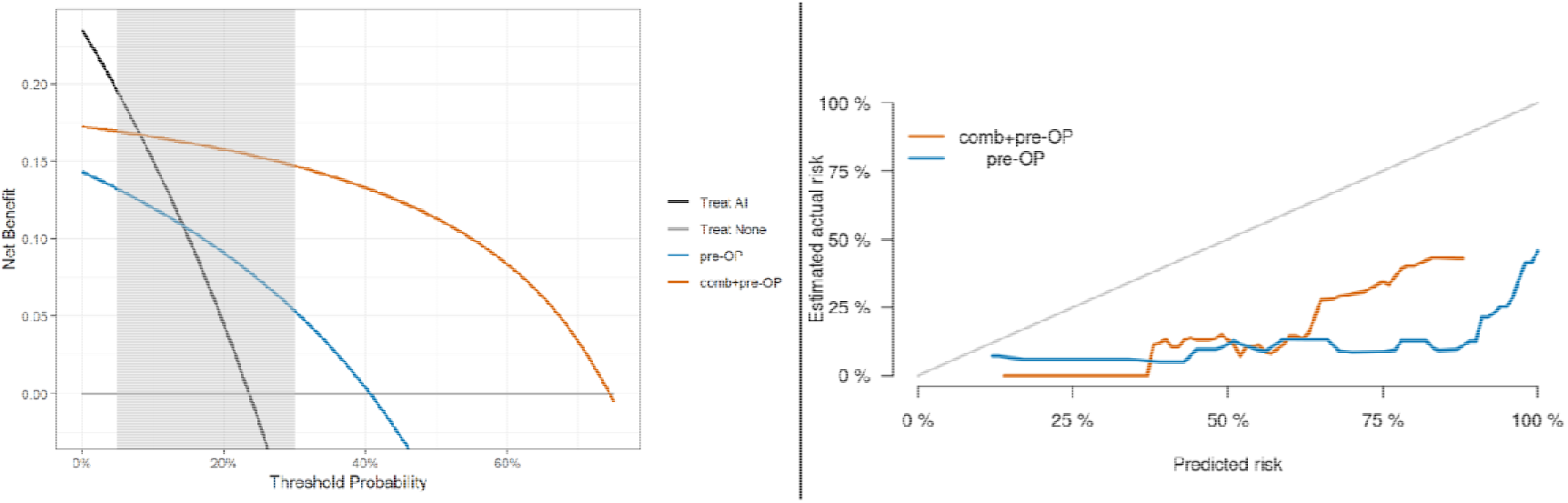
Decision curve analysis (left) and calibration curve (right). Using the same groups as in Figure 2, we found a net benefit of our predictive model compared to treating all patients in the relevant threshold range from five to 30% through decision curve analysis (left). A decision model shows a clinical benefit if the respective curve shows larger net benefit values than reference strategies. The combination of radiomic features derived from the *metastasis*, *edema*, and *clinical* features (*comb+pre-OP*) resulted in a higher net benefit compared to using only the clinical *Pre-OP* features and treating all patients or none. The calibration curve on the right was created by transforming the continuous risk rank predicted by the best *comb+pre-OP* ENR model (in orange) and by the clinical *pre-OP* ENR model (in blue) to event probabilities at 24 months. Although both models seem to overestimate the actual risk of our patients, the *comb+pre-OP* model predicts the risk closer to the actual risk.

### The relevance of brain metastasis volume

The predictions of our *comb+pre-OP* ENR model did weakly correlate with the cumulative BMV or BMV of the largest BM (Spearman’s rank correlation: r = 0.246 (p = 0.014) and 0.254 (p = 0.011), respectively).

While cumulative BMV alone was highly predictive in the test cohort, with a CI of 0.76 in a univariate Cox analysis, it only achieved a CI of 0.53 in internal validation. Using the BMV of only the largest BM increased the internal validation and external testing performance to 0.55 and 0.77, respectively. There was no significant difference in the BMV between the training and test cohort (p = 0.64, Wilcoxon rank sum test).

Stratifying our test set into small and large BMs by dividing the set at the median volume resulted in groups with three and 13 events, respectively. Our best model scored a CI of 0.58 and 0.78 in the respective groups. Interestingly, the model significantly risk-stratified the patients in the small BMV group, but not in the high BMV group (corresponding Kaplan-Meier analysis are depicted in Supplementary Figures 3 and 4).

When repeating the training and testing with the radiomic features extracted only from the biggest BM, the ENR learner was able to reach a CI of 0.75 (*comb+clinical*, Table 3). The results obtained by the RF model even surpassed our previously best model by 0.01 (CI = 0.78, Supplementary Table 4).

**Table 3:** Performance in the test set with automated U-Net segmentations and segmentations of only the largest metastasis.

| Group | Learner | T1-CE | FLAIR | comb | comb + pre-OP | comb + pre-OP + BMV |
| --- | --- | --- | --- | --- | --- | --- |
| Manual Segmentation | ENR | 0.76<br>(0.63-0.84) | 0.50<br>(NA-NA) | 0.69<br>(0.55-0.80) | **0.77**<br>(0.61-0.87) | 0.72<br>(0.57-0.82) |
| U-Net Segmentation | ENR | 0.68<br>(0.54-0.79) | 0.43<br>(0.27-0.58) | 0.64<br>(0.47-0.74) | 0.72<br>(0.54-0.82) | 0.67<br>(0.52-0.79) |
| largest BM | ENR | 0.75<br>(0.62-0.85) | 0.50<br>(NA-NA) | 0.65<br>(0.55-0.80) | 0.72<br>(0.60-0.86) | 0.73<br>(0.58-0.84) |

### End-to-end model using neural network-based automatic segmentations

To test the predictive value of neuronal network-based segmentations and therefore test the feasibility of a fully automated workflow, we conducted an additional parameter tuning and training run with radiomic features extracted from the automatically created segmentations. The results for our ENR learner are shown in Table 3. The best test results with this data were again obtained with the *comb+pre-OP* feature set (CI = 0.72). Overall, we observed an average decrease in performance by 0.06.

## Discussion

In this work, we were able to develop radiomics-based machine learning models that were able to predict FFLF better than clinical features alone. Our best model was trained with a combination of radiomic and clinical features and achieved a CI of 0.77 in a multicenter external test cohort outperforming any clinical predictive model. Our final model’s predictions significantly stratified the test patients into two risk groups and achieved an incremental net clinical benefit.

When using automatically generated segmentations from a previously trained neural network, the models performed slightly worse, with an average performance loss of 0.06. Still, the *comb+pre-OP* ENR model was able to reach a respectable CI of 0.72 in external testing. This demonstrates that an end-to-end solution is possible without clinician intervention.

The results in the external test cohort were, on average, better by a CI of 0.04. This may be explained by the larger amount of data available for training: The models tested on the external cohort were trained on all training data, while for internal validation, only 80% of the data was used for training, while testing was performed on the remaining 20%.

Several studies have approached predicting the LF of BMs. Most of them interpreted the prediction as a classification task and therefore only predicted whether an event occurred at a predetermined time^16–18,49–58^. In contrast, we approached the task as a survival task and therefore predicted a combination of event and time in terms of FFLF.

Another study predicting event and time of local failure by Huang *et al.*^59^ used Cox proportional hazards models and found that non-small cell lung cancer BMs with a higher zone percentage were more likely to respond favorably to Gamma Knife radiosurgery. In contrast to the treatment with surgery and adjuvant SRT in our study, the aforementioned studies focused on BMs treated with SRT, WBRT, and immune checkpoint inhibitors. Only one monocentric study with 67 patients by Mulford *et al.*^52^ investigated the prediction of local recurrence after surgical resection and adjuvant stereotactic radiosurgery, and found that radiomic features provided more robust predictive models of local control rates than clinical features (AUC = 0.73 vs. 0.40). Unlike our study, they predicted local failure as a binary classification task.

Another unique feature of our study is the multicenter external test cohort with patients treated at five different centers in multiple countries. In contrast to our study, the aforementioned studies all tested their models on an internal validation set and were therefore not tested on such a wide variety of scanners and imaging protocols as our models were.

Contrary to findings in previous studies^60^, the cumulative BMV and the BMV of the largest BM were not predictive in the internal validation, where they only reached a CI of 0.53 and 0.55, respectively. Since outcome and BMV appear to be independent in the training cohort, radiomic features representing BM size were not selected by our feature reduction algorithm. The only selected shape class feature in the best-performing feature set was metastasis flatness. Moreover, there was only a minor correlation (r = 0.25) between the predictions of the radiomic model and BMV. This shows that Radiomics can predict local failure based on features that do not directly represent BM size or volume.

Compared to approaches focusing on the use of neural networks, the use of classical machine learning has some advantages: Because only a small number of features are fed into the model, it becomes more comprehensible. Since it is known how the radiomics features are computed, it is possible to infer the clinical correlates. Neural networks, on the other hand, are more of intransparent black boxes, and it is difficult to understand exactly which characteristics of the tumor are predictive. In addition, neural networks often require the use of a graphics processing unit (GPU) to complete predictions in a reasonable amount of time, while our models run on the central processing unit (CPU) and can, therefore, run on low-end hardware.

Nevertheless, this work has several limitations: Training the models with only a limited number of features extracted from the segmentations prevents them from taking other factors into account, such as the surrounding tissue. Furthermore, segmentations of consistent quality are necessary for reliable results. In this study, all segmentations were created by the same person. To reduce the influence of the personal segmentation style, only features with a high correlation between manual and automatic segmentations were used for further modeling. The sole use of automatically generated segmentations may help with this limitation.

Around one-quarter of our patients had multiple BMs. By using the cumulative BMV as a feature, we not only took the volume of the resected BM into account but also the volume of all additional BMs. In our additional analysis, where we only considered radiomic features extracted from the largest metastasis, the best result improved slightly, while the mean across all models decreased by 0.01 compared to using the combined segmentation of all BMs. From this, we can conclude that segmenting all BMs did not harm the prediction of local failure of the resected BM.

In addition, radiomic features were extracted from a total of twelve synthesized T2-FLAIR sequences (six in the training cohort and six in the test cohort). Excluding these patients from the training and test sets resulted in a slight increase in performance. The largest increase in performance was found in the combined feature set (CI = 0.72 from 0.69). Furthermore, the *T1-CE* model showed the second largest increase in performance, surpassing our previous best feature set (*comb+pre-OP*), which showed no change in performance. Since the new best model did not even include features extracted from the T2-FLAIR sequence, we can conclude that radiomic features extracted from the synthesized T2-FLAIR sequences did not noticeably affect the performance of our model and the increase in performance may be attributed to the exclusion of difficult cases.

Despite these limitations, we were able to develop a model to predict freedom from local progression of BMs after resection and adjuvant SRT. The model performed well in a multicenter external test cohort with a variety of MRI scanners and imaging and therapy protocols. This model may help to tailor treatment to a patient’s individual risk of metastasis recurrence, thereby improving the overall management of BMs. We have published the model as an easy-to-use web app (https://jbuchner.shinyapps.io/shiny/), where the user can either upload the required MRI sequences and segmentations or input previously extracted radiomic features.

## Conflict of Interest

AW: Consultant: Gilead and Hologic Medicor GmbH; Honoraria: ACCURAY International, Universitätsklinikum Leipzig AöR, and Sanofi-Aventis GmbH; Board: IKF GmbH am Krankenhaus Nordwest

BeM: Grants: BrainLab, Zeiss, Ulrich, and Spineart; Royalties: Medacta and Spineart; Consultant and Honoraria: Medacta, Brainlab, and Zeiss; Stock: Sonovum

MG: President-Elect of ESTRO

NA: Independent Contractor: SAKK - Swiss Association for Clinical Cancer Research; Board: AstraZeneca; Research funding: ViewRay Inc.; Stock: Moderna Inc. and Idorsia AG; Chair: EORTC and Global Harmonization Group

SR: Honoraria: Brainlab

OB: Grants: European Union’s Horizon 2020 research and innovation programme; Board: working groups for Stereotactic Radiotherapy of the German Radiation Oncology and Medical Physics Societies, Section Editor of Strahlentherapie und Onkologie Journal

ALG: Research funding: Novocure

SEC: Honoraria and travel expenses: Roche, Bristol-Myers Squibb, Brainlab, AstraZeneca, Accuray, Dr. Sennewald, Daiichi Sankyo, Elekta, Medac, Icotec AG, HMG Systems Engineering, and Carl Zeiss Meditec AG

DB: Honoraria and travel expenses: Novocure

The remaining authors have no potential conflicts of interest to disclose.

## Authorship

All authors were involved in the data curation and acquisition of resources.

Formal analysis, methodology, and software: JAB, JCP

Visualization: JAB

Writing - Original Draft: JAB, FK, BW, JCP

Writing - Review & Editing: JAB, FK, SMC, TBB, AW, BeM, SR, OR, OB, CoZ, ABZ, ALG, BW, JCP

Supervision: MP, SEC, BW, JCP

Project administration: KAE, SEC, DB, JCP

Funding acquisition: SEC, RU, BW, JCP

All authors approved the manuscript.

## Data Availability

The trained model is available as a shiny web app. Training and test data are not publicly available.

