## Supplemental material for "Radiomics-based prediction of local control in patients with brain metastases following postoperative stereotactic radiotherapy"

Supplementary material

- Supplementary Table 1: Radiomic features and extraction parameter
- Supplementary Table 2: Radiomic feature sets
- Supplementary Table 3: Performance in internal validation and external testing
- Supplementary Table 4: Performance in external testing for all available data sets
- Supplementary Table 5: Beta values and normalization parameters
- Supplementary Figure 1: Time-dependent area under the receiver operating characteristic curve
- Supplementary Figure 2: Kaplan-Meier analysis with three risk groups
- Supplementary Figure 3: Kaplan-Meier analysis for small BMs
- Supplementary Figure 4: Kaplan-Meier analysis for large BMs

Supplementary Table 1: Radiomic features and extraction parameter

Image discretization was performed using a bin width of 10. Image intensity normalization was achieved via redistributing the image at the mean with a standard deviation and a scale of 100. Radiomic features were extracted using pyradiomics (version 3.01) in python (3.7). Following the Imaging Biomarker Standardization Initiative (IBSI) recommendations as previously described, a total of 104 features including first-order, shape, and texture features were extracted per sequence from the segmentation (shape features) or the original image. All extracted features are listed below.

|  | **Shape Features** |
| --- | --- |
| 1.) | Volume |
| 2.) | Surface Area |
| 3.) | Surface Volume Area |
| 4.) | Sphericity |
| 5.) | Spherical Disproportion |
| 6.) | Maximum 3D Diameter |
| 7.) | Maximum 2D Diameter Slice |
| 8.) | Maximum 2D Diameter Column |
| 9.) | Maximum 2D Diameter Row |
| 10.) | Major Axis |
| 11.) | Minor Axis |
| 12.) | Least Axis |
| 13.) | Elongation |
| 14.) | Flatness |
|  | **First Order Features** |
| 1.) | Energy |
| 2.) | Intensity Histogram Entropy |
| 3.) | Minimum |
| 4.) | 10th Percentile |
| 5.) | 90th Percentile |
| 6.) | Maximum |
| 7.) | Mean |
| 8.) | Median |
| 9.) | Interquartile Range |
| 10.) | Range |
| 11.) | Mean Absolute Deviation (MAD) |
| 12.) | Root Mean Squared (RMS) |
| 13.) | Skewness |
| 14.) | Excess Kurtosis |
| 15.) | Variance |
| 16.) | Intensity Histogram Uniformity |
|  | **Gray Level Co-occurrence Matrix (GLCM) Features** |
| 1.) | Autocorrelation |
| 2.) | Joint Average |
| 3.) | Cluster Prominence |
| 4.) | Cluster Shade |
| 5.) | Cluster Tendency |
| 6.) | Contrast |
| 7.) | Correlation |
| 8.) | Difference Average |
| 9.) | Difference Entropy |
| 10.) | Difference Variance |
| 11.) | Joint Energy (IBSI: Angular Second Moment) |
| 12.) | Joint Entropy |
| 13.) | Informal Measure of Correlation (IMC) 1 |
| 14.) | Informal Measure of Correlation (IMC) 2 |
| 15.) | Inverse Difference Moment (IDM) |
| 16.) | Inverse Difference Moment Normalized (IDMN) |
| 17.) | Inverse Difference (ID) |
| 18.) | Inverse Difference Normalized (IDN) |
| 19.) | Inverse Variance |
| 20.) | Maximum Probability (IBSI: Joint maximum) |
| 21.) | Sum Entropy |
| 22.) | Sum of Squares (IBSI: Sum of Squares) |
| 23.) | Maximal Correlation Coefficient (MCC) |
|  | **Gray Level Size Zone Matrix (GLSZM) Features** |
| 1.) | Small Area Emphasis (SAE) |
| 2.) | Large Area Emphasis (LAE) |
| 3.) | Gray Level Non-Uniformity (GLN) |
| 4.) | Gray Level Non-Uniformity Normalized (GLNN) |
| 5.) | Size-Zone Non-Uniformity (SZN) |
| 6.) | Size-Zone Non-Uniformity Normalized (SZNN) |
| 7.) | Zone Percentage (ZP) |
| 8.) | Gray Level Variance (GLV) |
| 9.) | Zone Variance (ZV) |
| 10.) | Zone Entropy (ZE) |
| 11.) | Low Gray Level Zone Emphasis (LGLZE) |
| 12.) | High Gray Level Zone Emphasis (HGLZE) |
| 13.) | Small Area Low Gray Level Emphasis (SALGLE) |
| 14.) | Small Area High Gray Level Emphasis (SAHGLE) |
| 15.) | Large Area Low Gray Level Emphasis (LALGLE) |
| 16.) | Large Area High Gray Level Emphasis (LAHGLE) |
|  | **Gray Level Run Length Matrix (GLRLM) Features** |
| 1.) | Short Run Emphasis (SRE) |
| 2.) | Long Run Emphasis (LRE) |
| 3.) | Gray Level Non-Uniformity (GLN) |
| 4.) | Gray Level Non-Uniformity Normalized (GLNN) |
| 5.) | Run Length Non-Uniformity (RLN) |
| 6.) | Run Length Non-Uniformity Normalized (RLNN) |
| 7.) | Run Percentage (RP) |
| 8.) | Gray Level Variance (GLV) |
| 9.) | Run Variance (RV) |
| 10.) | Run Entropy (RE) |
| 11.) | Low Gray Level Run Emphasis (LGLRE) |
| 12.) | High Gray Level Run Emphasis (HGLRE) |
| 13.) | Short Run Low Gray Level Emphasis (SRLGLE) |
| 14.) | Short Run High Gray Level Emphasis (SRHGLE) |
| 15.) | Long Run Low Gray Level Emphasis (LRLGLE) |
| 16.) | Long Run High Gray Level Emphasis (LRHGLE) |
|  | **Neighbouring Gray Tone Difference Matrix (NGTDM) Features** |
| 1.) | Coarseness |
| 2.) | Contrast |
| 3.) | Busyness |
| 4.) | Complexity |
| 5.) | Strength |
|  | **Gray Level Dependence Matrix (GLDM) Features** |
| 1.) | Small Dependence Emphasis (SDE) |
| 2.) | Large Dependence Emphasis (LDE) |
| 3.) | Gray Level Non-Uniformity (GLN) |
| 4.) | Dependence Non-Uniformity (DN) |
| 5.) | Dependence Non-Uniformity Normalized (DNN) |
| 6.) | Gray Level Variance (GLV) |
| 7.) | Dependence Variance (DV) |
| 8.) | Dependence Entropy (DE) |
| 9.) | Low Gray Level Emphasis (LGLE) |
| 10.) | High Gray Level Emphasis (HGLE) |
| 11.) | Small Dependence Low Gray Level Emphasis (SDLGLE) |
| 12.) | Small Dependence High Gray Level Emphasis (SDHGLE) |
| 13.) | Large Dependence Low Gray Level Emphasis (LDLGLE) |
| 14.) | Large Dependence High Gray Level Emphasis (LDHGLE) |

Supplementary Table 2: Radiomic feature sets

|  | **Radiomic Features** | | | |
| --- | --- | --- | --- | --- |
| **Feature set** | **Firstorder** | **Shape** | **GLCM** | **GLSZM** |
| **T1-CE** | Kurtosis,  Minimum  Skewness | Flatness | ClusterShade, MCC | ZoneEntropy |
| **FLAIR** |  | Elongation, Sphericity | ClusterShade |  |
| **comb** | Skewness (T1-CE), Kurtosis (T1-CE),  Minimum (T1-CE) | Flatness (T1-CE) | ClusterShade (T1-CE),  ClusterShade (FLAIR) | ZoneEntropy (T1-CE) |

We created multiple feature sets with 3-15 features per set with MRMR. In a nested CV, the best number of features per set was determined. This resulted in a *T1-CE*, *FLAIR*, and *comb* feature set with seven, three, and seven features, respectively. Six of the seven *comb* features were extracted from the metastasis segmentation from the T1-CE sequence, while only one feature was extracted from the edema segmentation from the FLAIR sequence.

Supplementary Table 3: Performance in internal validation and external testing

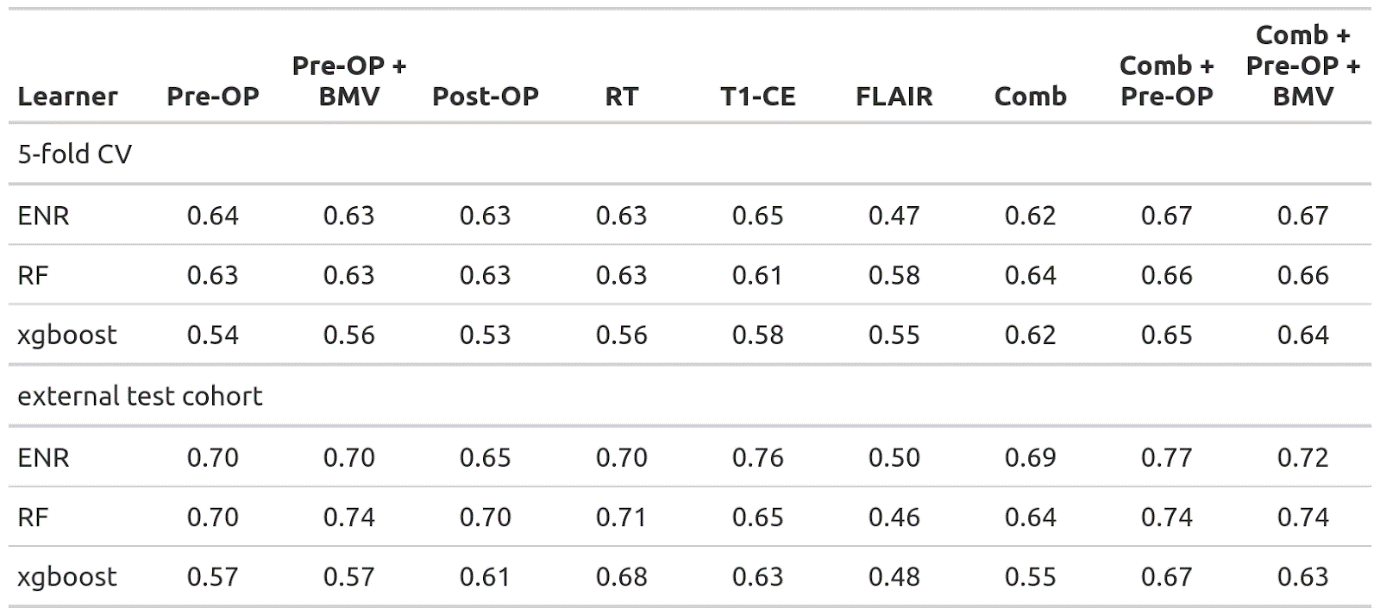

This is a supplement to Table 2: For our main model, we only used the learner that ranked highest in the internal validation (ENR). For completeness, we also tested the other two learners in the external test cohort. On average, RF performs slightly worse than ENR, with xgboost lagging behind.

Supplementary Table 4: Performance in external testing for all available data sets

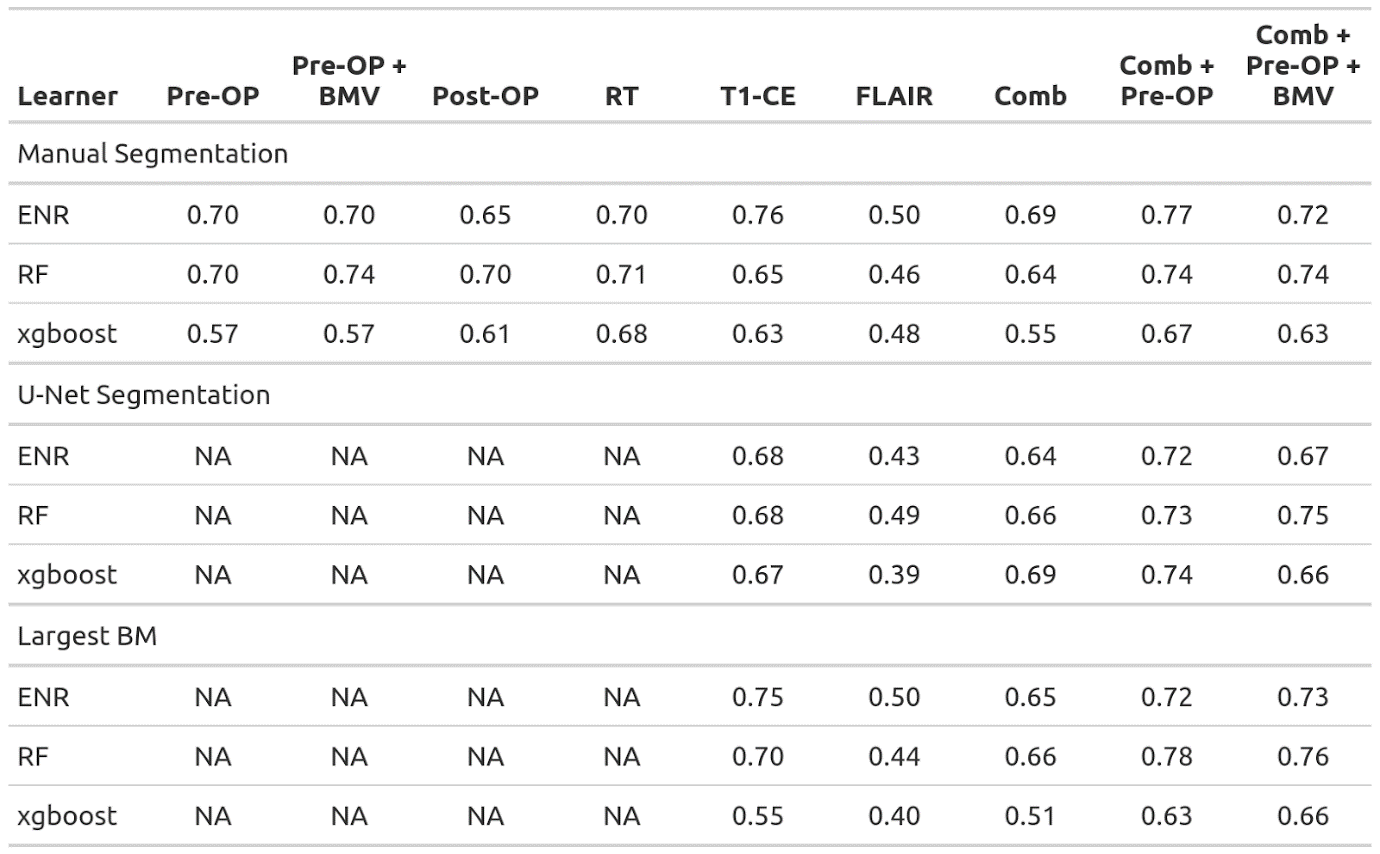

This is a supplement to Table 3: For completeness, we also tested xgboost and RF with the u-Net segmentations and segmentations of only the largest BM. The RF learner was able to surpass the previous best result with a CI of 0.78 achieved on the largest BM dataset with the *comb+pre-OP* feature set.

Supplementary Table 5: Beta values and normalization parameters

| **Type** | **Feature** | **Beta value** | **Normalization parameter** | | |
| --- | --- | --- | --- | --- | --- |
|  |  |  | **Mean** | **STD** | **Yeo-Johnson parameter** |
| **Radiomic feature** | Kurtosis (T1-CE) | 0.115430 | -0.04540 | 0.81238 | 0.10526 |
|  | Minimum (T1-CE) | -0.119190 | -0.09489 | 54.39253 | 0.85717 |
|  | Skewness (T1-CE) | 0.168315 | 0.31676 | 0.47683 | 0.72141 |
|  | Flatness (T1-CE) | 0.068694 | 0.61728 | 0.21753 |  |
|  | ClusterShade (T1-CE) | -0.020916 | 2555.12787 | 12146.61378 | 0.95920 |
|  | ClusterShade (FLAIR) | -0.039389 | -741.96326 | 3425.09835 | 0.9599 |
|  | ZoneEntropy (T1-CE) | 0.097210 | 7.21587 | 0.28464 |  |
| **Clinical feature (numeric)** | Age | -0.000793 |  |  |  |
|  | KPS | 0.004581 |  |  |  |
| **Clinical feature (location)** | Frontal | 0.150409 |  |  |  |
|  | Temporal | -0.438724 |  |  |  |
|  | Parietal | -0.146378 |  |  |  |
|  | Occipital | -0.413498 |  |  |  |
|  | Cerebellar | 0.250252 |  |  |  |
|  | Other | 0.944108 |  |  |  |
| **Clinical feature (histology of primary tumor)** | NSCLC – not further specified | 0 |  |  |  |
|  | NSCLC – non-adenocarcinoma | 0.535508 |  |  |  |
|  | NSCLC – adenocarcinoma | -0.496934 |  |  |  |
|  | Melanoma | 0.033393 |  |  |  |
|  | RCC | -0.079583 |  |  |  |
|  | Breast | 0.092171 |  |  |  |
|  | GI | 0.458709 |  |  |  |
|  | Other | -0.003596 |  |  |  |

The beta values extracted from the best learner (*comb+pre-OP* ENR) are listed in this table. Please note that the beta values do not directly correspond to feature importance since the variables are scaled differently. Furthermore, the radiomic features were normalized by z-score standardization and then applying the Yeo-Johnson transformation to transform the distribution of a variable into a Gaussian distribution. Therefore, these beta values cannot be applied to “raw” radiomic features. Therefore, we also supply the transformation parameters used for normalizing the radiomic features.

Supplementary Figure 1: Time-dependent area under the receiver operating characteristic curve

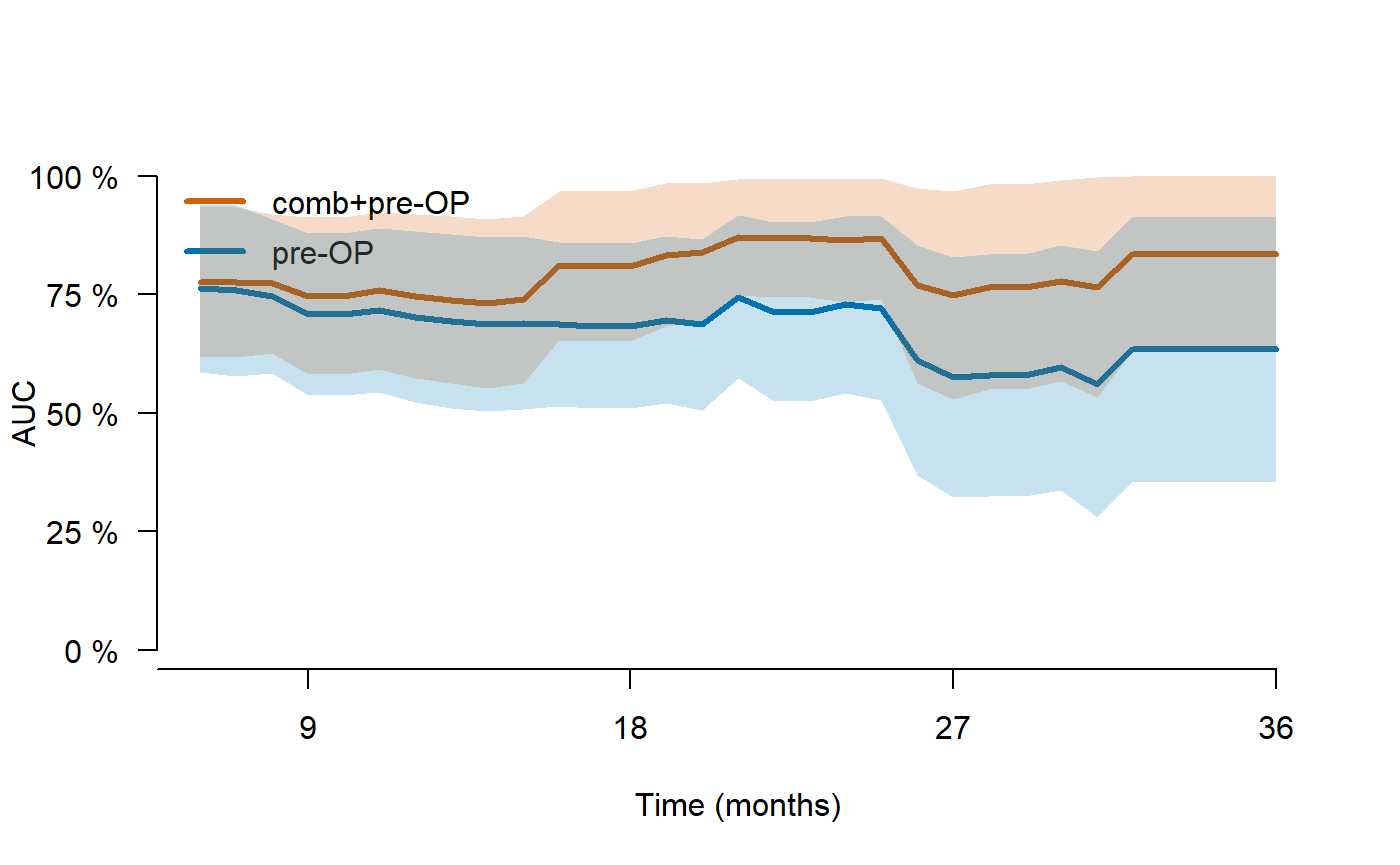

By converting the predicted continuous risk rank by our proposed *comb+pre-OP* ENR model and the clinical *pre-OP* ENR model to event probability distributions, we were able to calculate the area under the receiver operating characteristic curve (AUC) for different time points. In this figure, we plotted the time-dependent AUC for a range of follow-up times from six to 36 months. The mean AUC of our *comb+pre-OP* ENR model was 0.80 (orange) compared to 0.68 for the *pre-OP* model (blue).

Supplementary Figure 2: Kaplan-Meier analysis with three risk groups

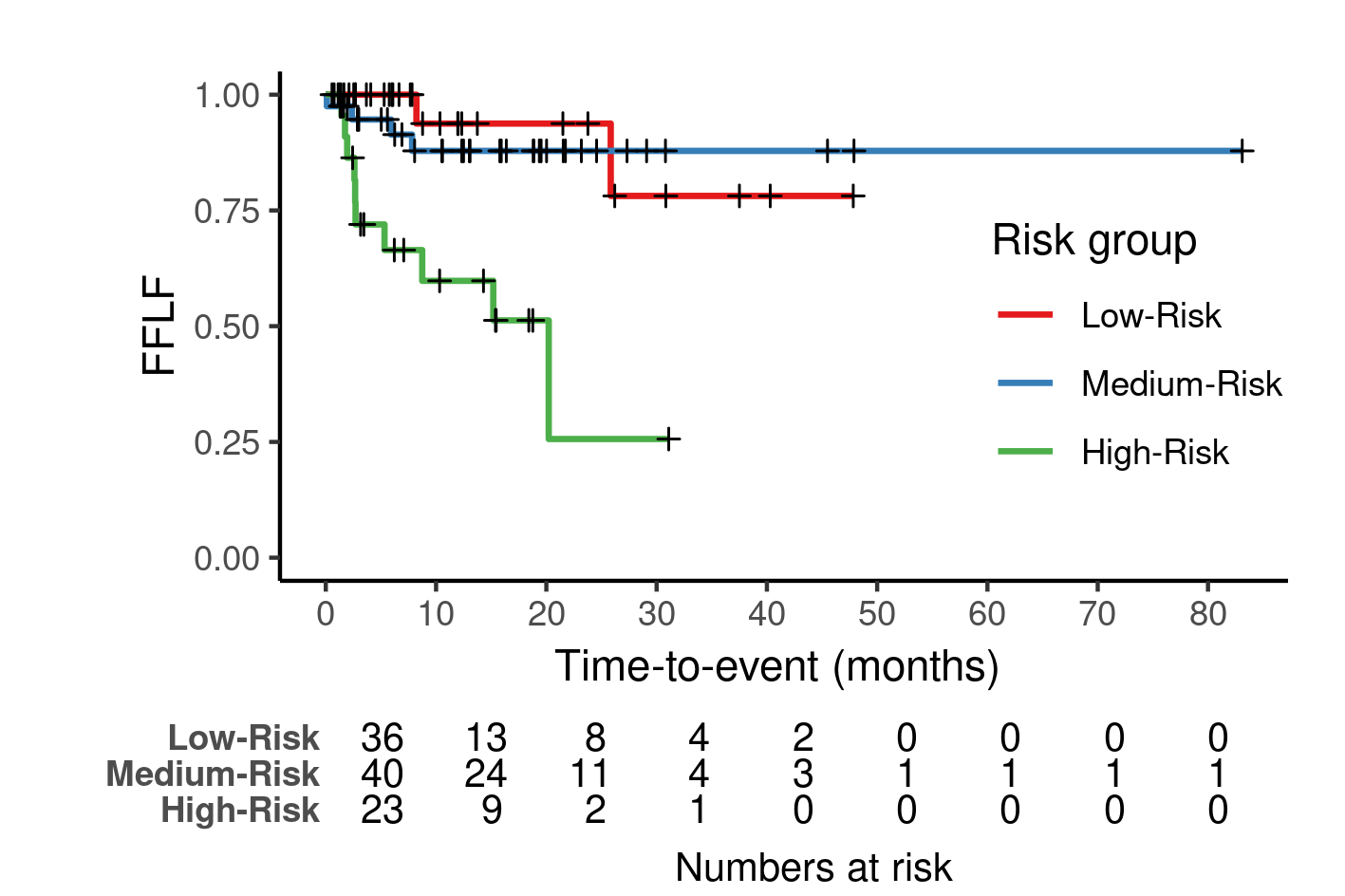

Using the 33rd and 66th percentiles of the continuous risk ranks in the training cohort as cutoffs, we stratified the patients in our external test cohort into three risk groups. There were significant differences in the freedom from local failure (FFLF) between the groups (p = 0.0001). Since both the low- and medium-risk groups seem to be very similar in survival, we combined them into one single group for further analysis.

Supplementary Figure 3: Kaplan-Meier analysis for small BMs

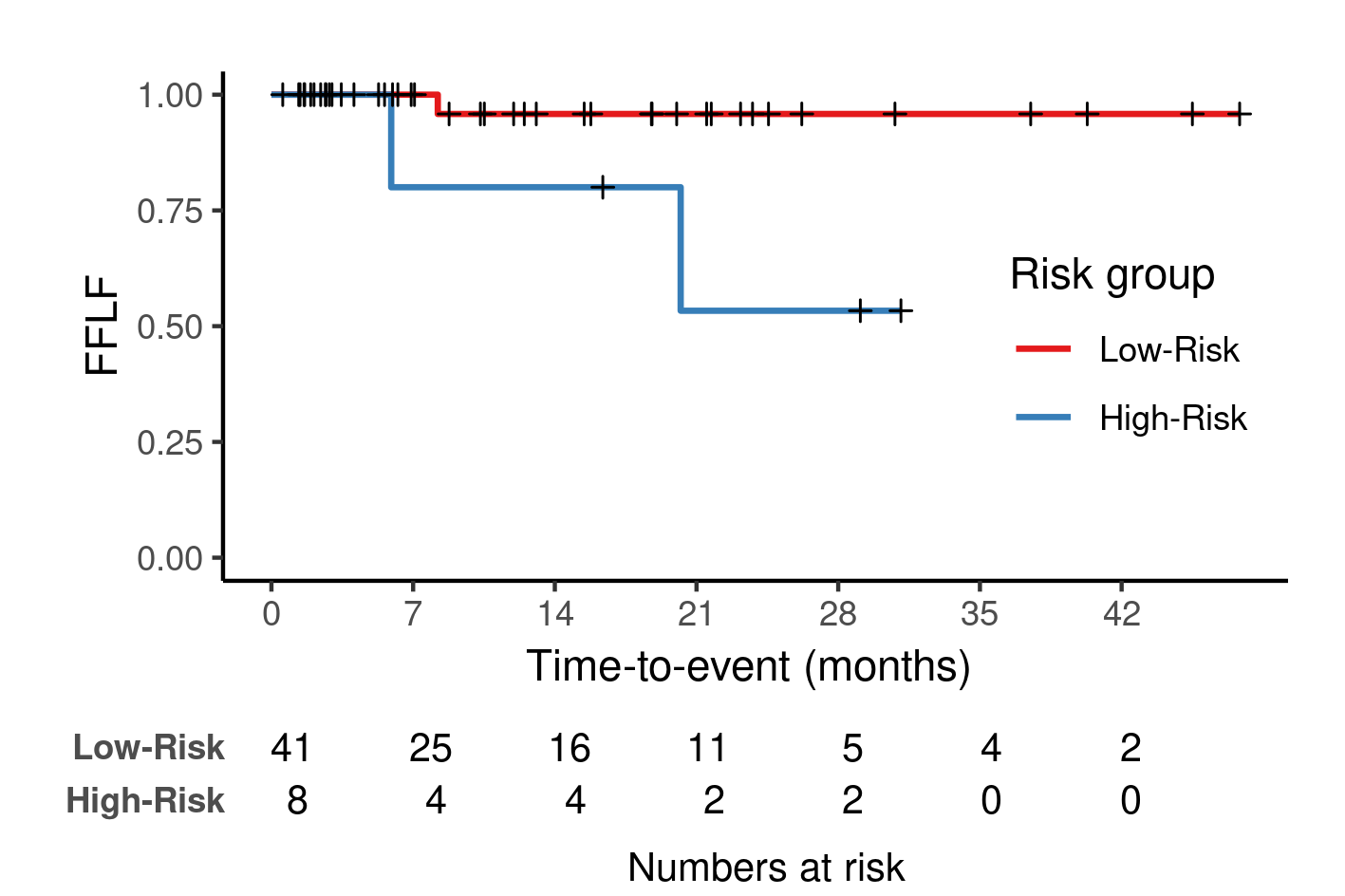

In this analysis, only patients with BMs smaller than the median size in the test set were included. In total, only three events occurred in this group. Even though the CI is relatively low (CI = 0.58), the model was able to significantly stratify the patients into two groups (p = 0.02) with the same cutoffs as in Table 2 (66th percentile of the continuous risk rank in the training cohort).

Supplementary Figure 4: Kaplan-Meier analysis for large BMs

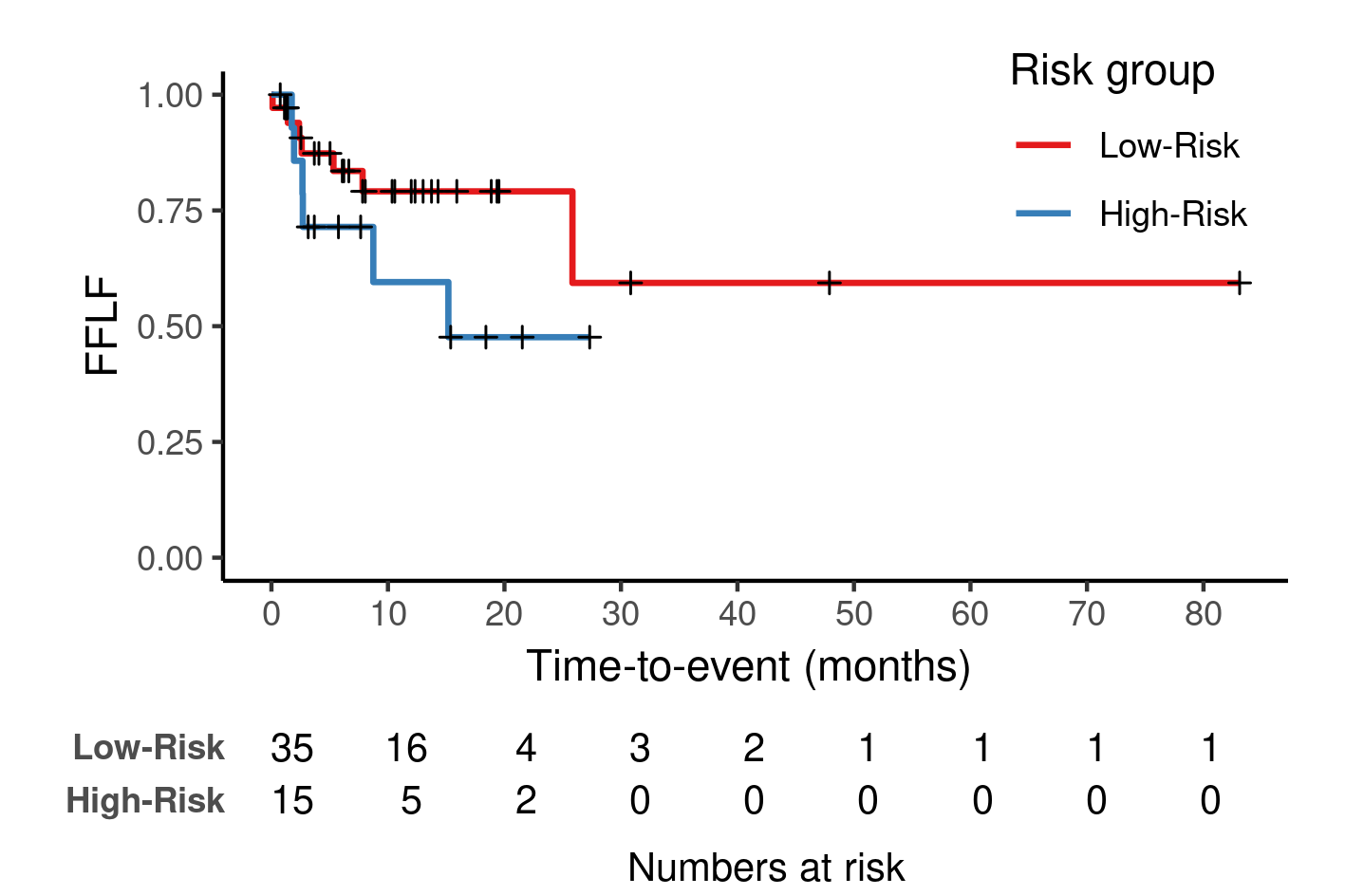

In this analysis, only patients with BMs larger than the median size in the test set were included. In total, 13 events occurred in this group. While the model reached a CI of 0.78, there was no significant difference between the two groups (p = 0.2).
